# Phase 2 Study Design and Analysis Approach for BBT-877: An Autotaxin Inhibitor Targeting Idiopathic Pulmonary Fibrosis

**DOI:** 10.1101/2025.03.31.25324971

**Authors:** Toby M. Maher, Jin Woo Song, Mordechai R. Kramer, Lisa Lancaster, Tamera J. Corte, Jeong H. Yun, KyungJin Kim, Jimin Cho, Luisa F. Sather, Peter M. George, Anand Devaraj, Jin Hyuk Jung, Sujin Jung

## Abstract

**Introduction:** Proof-of-concept (POC) studies are vital in determining the feasibility of further drug development, primarily by assessing preliminary efficacy signals with credible endpoints. However, traditional POC studies in idiopathic pulmonary fibrosis (IPF) can suffer from low credibility due to small sample sizes and short durations, leading to non-replicable results in larger Phase III trials. To address this, we are conducting a 24-week POC study with 120 patients with IPF, using a statistically supported sample size and incorporating exploratory computed tomography (CT)-based imaging biomarkers, to support decision making in the case of non-significant primary endpoint results. This approach aims to provide data to enable a robust decision-making process for advancing clinical development of BBT-877.

**Methods and analysis:** In this phase II, double-blind, placebo-controlled study, approximately 120 patients with IPF will be randomized in a 1:1 ratio to receive placebo or 200mg of BBT-877 twice daily over 24 weeks, with stratification according to background use of an antifibrotic treatment (pirfenidone background therapy, nintedanib background therapy or no background therapy). The primary endpoint is absolute change in FVC (mL) from baseline to week 24. Key secondary endpoints include change from baseline to week 24 in % predicted FVC, diffusing capacity of the lung for carbon monoxide, 6-minute walk test, patient-reported outcomes, pharmacokinetics and safety, and tolerability. Key exploratory endpoints include eLung-based CT evaluation and biomarker-based assessment of pharmacodynamics.

**Ethics and Dissemination:** This study is being conducted following the Declaration of Helsinki principles, Good Clinical Practice guidance, applicable local regulations, and local ethics committees. An independent data monitoring committee unblinded to individual subject treatment allocation will evaluate safety and efficacy data on a regular basis throughout the study. The results of this study will be presented at scientific conferences and peer-review publications.

**Trial Registration Number:** NCT05483907

**Key Message:** *What is already known on this topic:* Idiopathic pulmonary fibrosis (IPF) is characterized by progressive fibrosis, a high mortality rate and few effective treatment options. Proof of concept studies in IPF have not always translated into successful phase III clinical trials.

*What this study adds:* A description of the rationale, study design, methods, and analysis plan for a phase II, double-blind, placebo-controlled study of BBT-877 in patients with IPF, either alone or in addition to background antifibrotic treatment (nintedanib or pirfenidone).

*How this study might affect research, practice, or policy:* This phase II study design is expected to influence future research methodologies and clinical trial practices by providing a robust framework for studying IPF. Additionally, once the ongoing trial with BBT-877 is completed, the research methods will ensure credibility of the findings and will inform future clinical trial design.

## Introduction

Idiopathic pulmonary fibrosis (IPF) is a severe interstitial lung disease characterized by progressive scarring of the lung and a high mortality (1). The existing antifibrotic medications, pirfenidone and nintedanib, attenuate the rate of annual lung function decline in individuals with IPF by approximately 50% but do not halt the disease process or improve quality of life (2,3). Both drugs are associated with notable side effects which may be mitigated by dose reduction. Given the limited effectiveness of these currently approved treatments, there is an urgent need for novel therapeutic approaches.

Autotaxin (ATX), also known as ectonucleotide pyrophosphatase/phosphodiesterase family member 2, has lysophospholipase D activity that produces lysophosphatidic acid (LPA), a key lipid signaling molecule. LPA binds to specific receptors (LPARs) on target cells, mediating responses crucial to the development of conditions such as cancer and fibrosis by promoting cell motility, survival, and proliferation (4). Preclinical studies highlight the ATX-LPAR pathway as a promising target for treating pulmonary fibrosis, including IPF. Pharmacological inhibition of ATX and LPAR1 lead to decreased lung fibrosis, vascular leakage, and mortality while LPAR2 deficiency improves outcomes in rodent fibrosis models (5–7).

BBT-877, a potent ATX inhibitor (figure 1), has demonstrated strong inhibition of LPA-mediated chemotactic effects with low cytotoxicity in *in vitro* and *in vivo* studies (8). This Phase II proof of concept (POC) study seeks to address the unmet medical needs in IPF treatment by evaluating the potential of BBT-877 as a novel therapeutic option for individuals with IPF.

**Figure 1.**
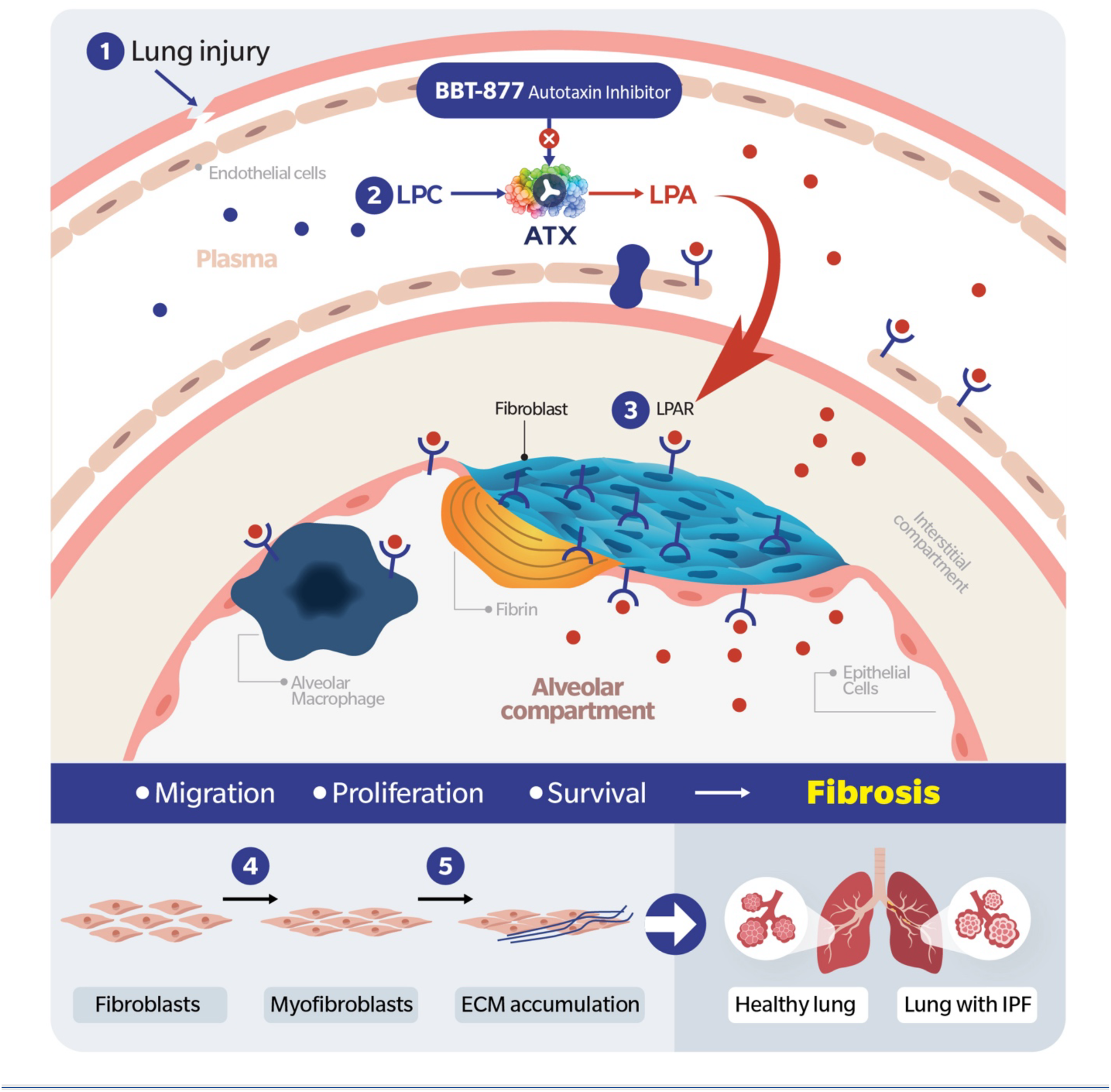
BBT-877 is a small-molecule compound that suppresses inflammation and fibrosis by reducing LPA production through the selective inhibition of autotaxin. Abbreviations: lysophosphatidylcholine (LPC); lysophosphatidic acid (LPA); autotaxin (ATX); lysophosphatidic acid receptor (LPAR); extracellular matrix (ECM); interstitial pulmonary fibrosis (IPF).

In the landscape of pharmaceutical development, POC studies serve as a crucial step in determining the feasibility of further clinical development programs. These studies are designed to assess preliminary efficacy signals through highly credible endpoints, providing a foundation for go/no-go decisions. The frequency of Phase III trial failures, despite promising results in earlier phases in IPF (9–12), underscores the importance of ensuring that efficacy results are replicable in larger sample sizes. Traditional Phase II studies, typically limited to 12 weeks and small sample sizes, often lack the statistical power to provide robustly reproducible efficacy results.

To address these challenges, we describe a 24-week Phase II POC study involving 120 patients with IPF. The study duration is 24 weeks, and the sample size is determined based on anticipated FVC change. Additionally, we have incorporated exploratory endpoints including AI-based CT biomarkers to add confidence to the go/no-go decision-making process and to hedge against potential variability of forced vital capacity (FVC) data. e-Lung (Brainomix, Oxford) imaging biomarkers, including the weighted reticulovascular score (WRVS), are generated for each CT scan by applying a validated AI-based convolutional neural network algorithm. e-Lung, has been associated with a higher likelihood of a relative decline in lung function of 10% over 52 weeks in IPF patients (12). In one study, WRVS has shown to change over time, with a difference of 3% being associated with a subsequent increased risk of death (13).

Our primary objective is to evaluate the efficacy of BBT-887 in individuals with a centrally adjudicated diagnosis of IPF. With a relatively extended duration of 24-weeks, an increased sample size based on statistical rationale, and a comprehensive set of secondary endpoints, this Phase II study aims to generate robust data to guide future development decisions.

This approach is expected to provide a range of complementary results that will increase the likelihood of ensuring a positive phase III program assuming success criteria are met.

## Methods and Analysis

### Study Design

This phase II, double-blind, placebo-controlled study is designed to evaluate the efficacy, safety and tolerability of BBT-877 in patients with IPF (NCT05483907). Approximately 120 patients will be randomized in a 1:1 ratio to receive placebo or 200mg of BBT-877 twice daily with the use of an interactive response system (IXRS). Randomization will be stratified into three groups according to background use of an antifibrotic treatment at screening (without antifibrotic background therapies, with pirfenidone background therapy, and with nintedanib background therapy). The study comprises an up to 6-week screening period; a 24-week treatment period; and a post-treatment follow-up period of 4-weeks (Figure 2).

**Figure 2.**
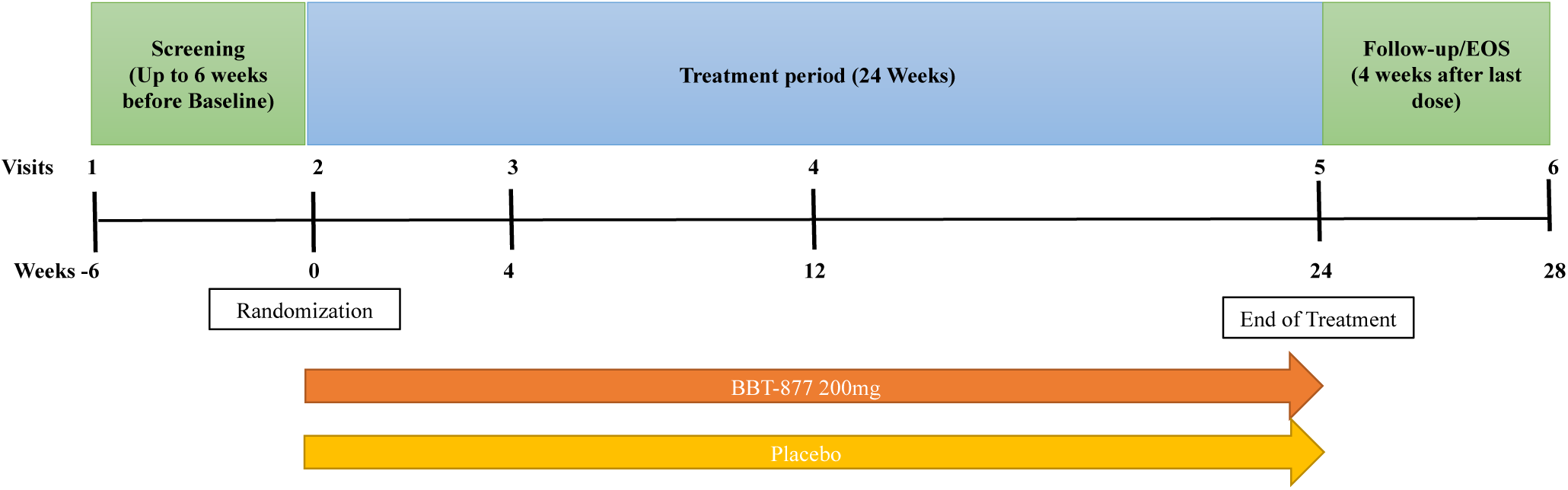
Study Design: Approximately 120 patients will be randomized in a 1:1 ratio to receive either placebo or 200mg of BBT-877 twice daily using an interactive response system (IXRS). Randomization will be stratified into three groups based on background antifibrotic therapy (no therapy, pirfenidone, or nintedanib). The study will include a 6-week screening period, a 24-week treatment period, and a 4-week post-treatment follow-up.

### Patient and Public Involvement

Patients and members of the public were not involved in the design, conduct, reporting, or dissemination plans of this research.

### Patient Population

Patient eligibility criteria are described in box 1. Patients must have a confirmed diagnosis of IPF by central review of HRCT imaging performed within 1 year, in accordance with 2018 ATS/ERS/JRS/ALAT clinical practice guideline (14). Patients with percent predicted forced vital capacity (ppFVC) ≥ 45%, a ratio of forced expiratory volume in the first second (FEV1) to FVC ≥ 0.7, and a diffusing capacity of the lung for carbon monoxide (DLco) corrected for hemoglobin ≥ 30% predicted will be eligible to participate. Male patients will have completed family planning and understood the potential risk of testicular toxicity. Women of childbearing potential will have a negative serum pregnancy test before treatment. Both male and female patients must use contraception throughout the study.

Patients with background use of antifibrotic treatment will be defined as patients on pirfenidone or nintedanib for at least 3 months and who are on a stable dose in the 4 weeks prior to screening. Patients without background use of antifibrotic treatment will be either treatment naive or have discontinued pirfenidone or nintedanib for at least 1 month prior to screening.

### Objectives and Endpoints

The primary endpoint will be absolute change in FVC (mL) from baseline to week 24. Spirometry measures will be assessed by spirometers (ERT SpiroSphere) provided by the sponsor, and reviewed centrally in real time by Clario throughout the study according to 2019 ATS/ERS Guideline (15) to ensure the quality of the data. This guideline includes the BEV standard as an acceptability criterion for both FEV1 and FVC, however the study will waive the BEV criterion for FVC acceptability. Quality of FEV1 and FVC of each effort will be determined and unacceptable efforts will be deselected.

Key secondary endpoints will include change from baseline to week 24 in % predicted FVC, DLco, distance assessed by 6-minute walk test (6MWT), patient-reported outcomes, and safety and tolerability. Pharmacokinetics (PK) will be also part of the secondary endpoints.

A key exploratory endpoint will be e-Lung assessment of baseline and follow up HRCT scans. Sites will be instructed to provide scans of slice thickness ≤ 2mm and a reconstruction increment ≤ 2mm, in accordance with study HRCT reconstruction parameters. We will evaluate baseline CT WRVS levels and compare means values across trial arms to exclude chance imbalances. We will investigate whether including baseline WRVS as a covariate improves precision, and therefore the power to detect a significant difference between groups. This will be quantified by measuring the R^2^ statistic, with and without WRVS in the model (16).

Finally, we will test the efficacy of BBT-877 on e-Lung biomarkers of disease severity including WRVS, e-Lung volume, ground glass opacification and total disease extent, using similar methods as used for ANCOVA analysis of the primary outcome, and adjusting for the baseline characteristics age, sex, baseline FVC, and treatment group. The WRVS is a % score which quantifies the extent of reticulovascular structures in the lung periphery. Reticulovascular structures are detected by an automated e-Lung algorithm that segments high-density elongated structures within the lung that have a shape and size typical of vessels, honeycombing walls and reticular patterns. These patterns can then be quantified using a weighting based on peripheral volume. The e-Lung volume is the total volume (in mL) of both lungs as measured by automated e-Lung segmentation algorithms. The ground glass opacification score is quantified in % estimating the proportion of the lung parenchyma affected by non-subtle elevations in density typical of ground glass opacification. The total disease extent biomarker quantifies a composite of the totality of reticulovascular structures and ground glass opacification across both lung fields.

Further exploratory analyses will include serial assessment of plasma LPA levels at baseline, week 4 and week 24. Plasma concentrations of LPA 18:2 will be determined using LC-MS/MS method qualified with respect to accuracy, precision, linearity, sensitivity, and specificity, with the analytical range of 0.5 −100 ng/mL, and the percent change of LPA 18:2 at each time point will be subsequently determined from the baseline sample. All endpoints are described in box 2.

### Statistical Analyses

The full analysis set (FAS) will consist of all randomized patients receiving at least 1 dose of study treatment and will be used for the primary efficacy analysis. The primary analysis will use a mixed model repeated measures (MMRM) model that includes the change from baseline to each visit in FVC as the dependent variable, the baseline FVC, sex, and age as covariates, and treatment, visit, the presence/absence of background therapy, and their 2- and 3-way interactions as fixed effects. No imputation will be performed for the primary analysis.

The sensitivity analyses will consider assumptions about missing FVC data at the Week 24 visit. A supportive analysis will use an analysis of covariance (ANCOVA) model that includes the change from baseline to Week 24 visit in FVC as the dependent variable, the baseline FVC as a covariate, and treatment and the presence/absence of background therapy as fixed effects. The second supportive analysis will use the same model as the primary efficacy analysis but will be based on the actual background therapy received for majority of the time, to investigate the impact of changes in the background therapy. Significance testing will be performed using a two-sided approach and 90% confidence interval will be provided.

To detect a 100mL difference in absolute change in FVC from baseline to week 24 between the BBT-877 treatment group and placebo groups, a sample size of 120 patients (60 per arm) is required. This sample size will provide 80% power to achieve statistical significance, assuming a standard deviation of 220mL (17).

### Study Governance

There are no planned efficacy interim analyses. An independent data monitoring committee (IDMC), unblinded to treatment allocation, will regularly evaluate safety and efficacy data throughout the study. The IDMC will provide recommendations to Bridge Biotherapeutics on whether the study should be continued, modified, or stopped.

## Discussion

### Rationale for Conducting the Study

Pirfenidone and nintedanib are the approved antifibrotic therapies for IPF, known to slow disease progression. Nevertheless, these two antifibrotics do not halt, much less reverse disease and can be limited due to frequently associated gastrointestinal and skin-related adverse events (2,18).

This is the first reported phase II study of BBT-877 in patients with IPF. It is known that ATX generates LPA which induces various cellular responses, including proliferation, inflammation, fibrosis, and migration (19). LPAR1 and 2 deficient mice actually showed protective effects against bleomycin-induced fibrosis, suggesting that inhibition of ATX may be a more effective way of targeting LPA-LPAR signal axis (20,21,7). Persistence of myofibroblast is one of the key features of IPF pathogenesis, and LPA-LPAR1 signaling promotes resistance to fibroblast apoptosis. In addition, recent evidence suggests that LPA-LPAR5 deactivates CD8 T cells and inhibits their migration and that this is another potential mechanism by which LPA-LPARs drive myofibroblast persistence (22–26). The important role for the LPA-Autotaxin axis in the pathogenesis of fibrosis supports the potential of BBT-877 as a potent ATX inhibitor to be an effective treatment for IPF. Although phase III trials of ziritaxestat, another ATX inhibitor, were terminated early due to its failure to improve clinical outcomes in patients (9), pre-clinical studies show that BBT-877 has a favorable potency and safety profile compared to ziritaxestat (8).

### Rationale for study design

The study population mirrors previous and ongoing IPF clinical trials. In some regards the study is even more inclusive of a broader population that better represents a real-world population of IPF patients with no upper limit in age, no restriction with regards background antifibrotic therapy and no exclusion of subjects on the lung transplant waiting lists. Antifibrotic drug modification, interruption or discontinuation will be allowed per investigator discretion. Initiation of antifibrotics will also be permitted at the discretion of the investigator during the study. As noted in recent FDA and investigator discussions, FVC has been emphasized as a crucial primary efficacy endpoint due to its biological plausibility and its effectiveness as a surrogate measure for mortality. A number of drugs tested in 12-week FVC studies have subsequently gone on to fail in late phase trials (27). It was therefore felt that a 24-week study should yield a richer dataset, enhancing the overall robustness when interpreting the final study results.

A 1:1 randomization ratio is adopted, as it is the most efficient and ensures the greatest power to detect differences between the two treatment groups (28).

### Rationale for dose selection

BBT-877 at a 200mg twice daily (BID) dose will be selected based on results from the Phase I first-in-human study, which included single-ascending doses (50 to 800mg) and multiple-ascending doses administered once daily (200mg to 800mg) and BID (100mg to 200mg) (NCT03830125). All dose levels were safe and well tolerated. Plasma LPA (18:2) inhibition was shown to be over 80% at both 100mg and 200mg BID (8). Consequently, a 200mg BID dose was deemed reasonable, with the option to reduce the dose to 100mg BID for safety reasons, while still maintaining effective exposure at steady state for plasma LPA inhibition.

### Rationale for endpoints

The primary efficacy endpoint of this study will be the change in absolute FVC (mL) from baseline to week 24. Change in FVC is highly predictive of outcome and prognosis, as demonstrated by the correlation between treatment effects on FVC and on mortality. Therefore change in FVC has been commonly used as the primary endpoint in IPF trials and has been used to gain approval of both nintedanib and pirfenidone (27). Patients in this study will have several FVC assessments performed including baseline, week 4, week 12 and week 24.

The secondary efficacy endpoints will include other methods of assessing the physiological severity of lung disease to be predictive of mortality risk, including change in DLco and 6MWT from baseline to week 24. Additional measures of efficacy will include Patient-Reported Outcomes (PROs) to evaluate changes in symptoms and their impacts on patients’ quality of life throughout the study. Utilizing tools including St. George’s Hospital Respiratory Questionnaire (SGRQ) (29), Living with Idiopathic Pulmonary Fibrosis Symptoms (L-IPF Symptoms) (30), Living with Idiopathic Pulmonary Fibrosis Impacts (I-IPF Impacts) (30), and Leicester Cough Questionnaire (LCQ) (31), will allow for a comprehensive understanding of how patients feel, ensuring that their experiences and perspectives are integral to the assessment of treatment effectiveness 6MWT and PROs will provide additional clinically meaningful measures of symptoms and functions which are not fully captured by change in FVC.

PK and PD will involve measuring pre-dose and 4-hour post-dose plasma concentrations of BBT-877, both alone and in combination with pirfenidone and nintedanib, respectively. This approach will allow for an evaluation of abbreviated PK and PD of BBT-877, providing insights into its pharmacological characteristics. It will also facilitate a preliminary assessment of potential drug-drug interactions (DDI).

Computerized evaluation of HRCT imaging at baseline and the end of study applying e-Lung biomarker outputs (WRVS, e-Lung volume, ground glass opacification and total disease extent) will be part of exploratory analyses, to study association with FVC decline and enrich patient recruitment in future studies. The data generated will be used to support any decision to take BBT-877 into later phase trials and furthermore will enable assessment of the relationship between baseline imaging characteristics and subsequent FVC decline. This might enable imaging-based enrichment strategies to be adopted in future studies.

## Conclusions

BBT-877 is an ATX inhibitor currently under clinical evaluation for the treatment of IPF. This drug is being assessed in a Phase II study involving patients with IPF. As with other previous Phase II studies in patients with IPF, this trial will evaluate efficacy with FVC as the primary objective. Patients will be allowed to continue receiving background antifibrotic therapy, mirroring real-world treatment scenarios. Additionally, this trial aims to generate further clinical evidence on the effectiveness of ATX inhibition as a treatment approach for IPF. It will include the generation of comprehensive PK and PD data, including the impact of treatment on LPA levels and the integration of imaging biomarkers using e-Lung, a novel quantitative CT based AI algorithm. This information is crucial for understanding the relationship between pharmacological effects and clinical outcomes. By providing these insights, the trial has the potential to offer a new, effective, and well-tolerated therapeutic option for patients with IPF. The findings from this study will play a critical role in shaping the next steps in our clinical development strategy. Specifically, they will guide the selection of endpoints and the determination of optimal sample sizes for future pivotal trials.

## Data Availability

This is a study-design manuscript and no data was analyzed or discussed for this manuscript.

## Author Contributorship Statement

This study was a collaborative effort involving multiple authors, each contributing uniquely to its success. Toby M. Maher, Sujin Jung, Jeong H. Yun, and KyungJin Kim were responsible for the conceptualization and design of the study. KyungJin Kim, Sujin Jung, Jimin Cho, Peter M. George, and Anand Devaraj contributed to the development of the data collection and analysis plan. The initial draft of the manuscript, along with subsequent revisions, was prepared by Toby M. Maher, KyungJin Kim, Jimin Cho, Luisa F. Sather, Sujin Jung, Jin Hyuk Jung, Peter M. George, and Anand Devaraj. Critical review and editing for intellectual content were provided by Toby M. Maher, KyungJin Kim, Sujin Jung, Luisa F. Sather, Jimin Cho, Jin Hyuk Jung, Jin Woo Song, Mordechai R. Kramer, Lisa Lancaster, Tamera J. Corte, and Jeong H. Yun. Sujin Jung supervised the study design, provided guidance and feedback, and approved the final version. Funding for the research was secured by Bridge Biotherapeutics, Inc., which also facilitated necessary resources. All authors reviewed and approved the final version of the manuscript for publication and agree to take responsibility for all aspects of the work.

## Acknowledgments

We extend our sincere gratitude to all investigators, site staff, and Bridge Biotherapeutics personnel, whose dedication and expertise were instrumental in designing this study.

## Patient Consent for Publication

Not required.

### Box 1.

Key Patient Eligibility Criteria

#### Inclusion Criteria

All patients

▪ Age > 40 years old.
▪ Diagnosis of IPF in accordance with ATS/ERS/JRS/ALAT guidelines.
▪ Chest HRCT performed within 12 months for IPF diagnosis by central review based on HRCT and lung biopsy.
▪ Able to walk at least 150 m during the 6MWT.
▪ Forced vital capacity ≥45% predicted.
▪ Ratio of forced expiratory volume in the first second (FEV1) to FVC ≥0.7.
▪ Diffusing capacity for the DLCO corrected for hemoglobin ≥30% predicted.
▪ Absence of IPF improvement in the past year.
▪ Patients receiving either pirfenidone or nintedanib, should be on it for at least 3 months and with stable dose in the 4 weeks prior to screening, OR taking neither pirfenidone nor nintedanib. If the patients were on pirfenidone or nintedanib previously, they should have been off for at least 1 month prior to screening.
▪ Agree to use contraception methods

Male patients:

▪ Have completed family planning, understand the risks of potential irreversible testicular toxicity and agree to participate.

#### Exclusion Criteria

▪ Unable to perform spirometry as per ATS.
▪ Evidence of IPF exacerbation within 3 months.
▪ Evidence of emphysema extent greater than the extent of fibrosis.
▪ History of lung transplant or lung volume reduction surgery.
▪ Current immunosuppressive condition.
▪ Congestive heart failure class III or IV according to New-York Heart Association classification.
▪ Pulmonary hypertension (PH) requiring PH specific therapy.
▪ Unstable cardiovascular, pulmonary or other disease within 6 months.
▪ Lower respiratory tract infection requiring antibiotics within 4 weeks.
▪ Interstitial lung disease associated with known primary diseases, exposures, and drugs.
▪ History of other types of respiratory diseases.
▪ History of malignancy within the past 5 years.
▪ Underwent major surgery within 3 months.
▪ Patients unable to refrain use of bronchodilator before assessments.
▪ Use any of the following therapies within 4 weeks before screening and during screening, or planned during the study:

- Endothelin receptor antagonists.
- Phosphodiesterase type 5 (PDE 5) inhibitors.
- Prednisone at steady dose >10mg/day or equivalent.
- Strong cytochrome P450 isoenzyme 3A4 (CYP3A4) and/or P glycoprotein (P-gp) inhibitor.

### Box 2.

Key Endpoints

Primary Endpoint:

- Change from baseline in FVC (in mL) compared to placebo at Week 24 stratified by presence/absence of background therapy (SoC).

Secondary Endpoints:

- Change from baseline in FVC % predicted compared to placebo at Week 24 stratified by presence/absence of background therapy (SoC).
- Change from baseline compared to placebo in DLCO at Week 24.
- Change from baseline in functional exercise capacity as measured by change in 6 minute walk distance assessed by the 6MWT at Week 24, compared to placebo.

